# Impact of electronic medical records on healthcare delivery in Nigeria: A Review

**DOI:** 10.1101/2023.12.05.23299498

**Authors:** Oreoluwa Olukorode, Oluwakorede Joshua Adedeji, Adetayo Adetokun, Ajibola Ibraheem Abioye

## Abstract

Electronic medical records (EMRs) have great potential to improve healthcare processes and outcomes. They are increasingly available in Nigeria, as in many developing countries. The impact of their introduction has not been well studied. We sought to synthesize the evidence from primary studies of the effect of EMRs on data quality, patient-relevant outcomes and patient satisfaction. We identified and examined five original research articles published up to May 2023 in the following medical literature databases: PUBMED/Medline, EMBASE, Web of Science, African Journals Online and Google Scholar. Four studies examined the influence of the introduction of or improvements in the EMR on data collection and documentation. The pooled percentage difference in data quality after introducing or improving the EMR was 142% (95% CI: 82% to 203%, *p-*value < 0.001). There was limited heterogeneity in the estimates (I^2^ = 0%, *p-* heterogeneity = 0.93) and no evidence suggestive of publication bias. The 5^th^ study assessed patient satisfaction with pharmacy services following the introduction of the EMR but neither had a comparison group nor assessed patient satisfaction before EMR was introduced. We conclude that the introduction of EMR in Nigerian healthcare facilities meaningfully increased the quality of the data.

**Author Summary:** Electronic medical records, or EMRs, offer the potential to improve healthcare in many developing nations, including Nigeria. The actual impact of these digital records hasn’t gotten enough attention, despite their growing availability. Our goal in doing this review was to compile and evaluate primary study data on the effects of EMRs on patient satisfaction, patient-related outcomes, and data quality in Nigeria. After searching through medical literature databases, we found five original research publications up to May 2023 that were suitable for our study. The impact of EMR implementation or upgrades on data collection and documentation was examined in four of these studies, while patient satisfaction with pharmacy services following EMR adoption was evaluated in the fifth study. Although the included studies did not quantitatively assess the impact of EMR adoption on patient satisfaction, we were able to conclude from the studies that the introduction of EMR in Nigerian healthcare facilities meaningfully increased the quality of the data

## Introduction

Electronic medical records (EMRs) are extensively used in developed countries but their adoption in developing countries has been slow [1]. According to a survey conducted by the World Health Organization, 114 countries are presently developing national electronic medical record systems due to an increasing awareness that a more robust health information technology (HIT) system is essential for delivering superior healthcare at reduced expenses [2]. The adoption of EMR in these countries has been hindered by implementation, maintenance, and lack of required computer skills among healthcare professionals [3].

The impact of the introduction of the EMRs has not been well studied. Most of the published literature have been perspective papers and reviews [4–6], case studies describing the practical experiences of hospitals setting up EMRs [7–10], primary studies exploring the barriers hindering their adoption [1,11–15] and studies leveraging EMRs for research [16–19]. Several primary studies have however assessed the impact of EMRs on patient-relevant outcomes [8,20– 25], but no reviews have synthesized the evidence from them. The magnitude or direction of the impact of EMRs on these outcomes therefore remains unclear.

The main objective of this review is to assess the impact of the introduction and use of EMRs in healthcare facilities in Nigeria on patient-relevant outcomes. We will thus synthesize the evidence from studies of patient waiting time, patient satisfaction and health outcomes.

## Methods

The guidelines from the Preferred Reporting Items for Meta-analysis, and Systematic Reviews (PRISMA) guidelines were used in the design, analysis, and reporting of this study [26].

### Systematic Search

We conducted a systematic search of medical literature databases in May 2023. The search included PubMed/Medline (U.S. National Library of Medicine), Embase (Elsevier), Web of Science and African Journals Online (AJOL). The search was based on a combination of keywords and Medical Subject Heading (MeSH) terms denoting “medical records” and “Nigeria” were used. The search was not restricted by year of publication, study design or by language. Hand-searching of the reference lists of included articles, related review papers, and the first 500 Google Scholar search hits were also reviewed for additional relevant papers.

### Selection of studies

Duplicate citations were removed and the title and abstract of identified studies were reviewed by at least two researchers working independently (OO, OA and AA). Discrepancies were resolved by a third author (AIA). The full text of qualifying articles were then reviewed in a similar manner. To be eligible for inclusion, studies had to be published in English Language, have assessed the impact of Electronic medical records, and conducted in Nigeria. Studies were excluded if they were systematic reviews, cross-sectional studies, case reports, and duplicate studies. Rayyan.ai was used to manage study selection [27].

### Data extraction

Data were extracted from the eligible studies using a data extraction sheet prepared by investigators. Data on study designs, population characteristics, covariates, study details, measures of association and their corresponding confidence intervals (CI), standard errors or *p*-values were extracted from the included studies.

### Study quality and risk of bias

Included studies were critically appraised for the risk of bias. The risk of bias was assessed using the Cochrane Collaboration’s revised tools for assessing the risk of bias in non-randomized studies [28]. No inclusion or analysis decisions were made based on the risk of bias.

### Meta-analysis

The ratio of the percentage in data quality was estimated and standard errors obtained from p-values[29]. Pooled estimates of the log ratio of means for data quality were obtained from random effects meta-analysis based on the restricted maximum likelihood since ≥ 3 studies reported estimates for the same intervention-outcome relationship [30]. The random effects approach was selected *a priori* because it accounts for variability in the true intervention effect [31]. The log ratio of means is a metric of the percentage difference and was preferred to the weighted and standardized mean difference measures as it is easily interpretable [32]. Heterogeneity was assessed using the I^2^ statistic to explore the degree of variability between the studies and explored in meta-regression. I^2^ was regarded as minimal if 0-40%, moderate if >40-60%, substantial if 60– 80% and considerable if >80% [33]. Publication bias was evaluated by visual inspection of funnel plots and the Egger’s test [34]. Meta-analysis was conducted using the Metafor package [35] in RStudio [36] *P*-values are 2-sided, and significance was set at *p-*value < 0.05.

## Results

Summary of included studies

We found five studies (**Fig 1**) that specifically analyzed the influence of EMR on healthcare outcomes in Nigerian hospitals. There was one cross-sectional study [23] and four longitudinal studies [20–22,24]. These studies were all conducted in public secondary and tertiary hospitals with the sample population ranging from doctors and nurses to patients. The risk of bias in the studies were low risk for two studies[20,21], some concern for one study [22], and high risk for two studies[23,24]. The search details are shown in Table 1 with the summary of included studies shown in Table 2.

**Table 1:** Search Details.

| Database | Search terms | Hits – 5<sup>th</sup><br>May, 2023 |
| --- | --- | --- |
| PubMed | ((((electronic[Text] OR digital[Text]) AND (medical records[Text] OR health records[Text])) OR ("medical records systems, computerized"[MeSH Terms])) AND (Nigeria[Text] OR Nigeria[Mesh])) | 47 |
| Embase | ('electronic medical record'/exp OR 'electronic medical record') AND ('nigeria'/exp OR Nigeria OR 'nigerian'/exp OR Nigerian) | 79 |
| Web of Science | (electronic health records (Title) or electronic health records (Abstract)) AND (Nigeria (Title) or Nigeria (Abstract)) | 40 |
| Google Scholar | ("electronic medical records" OR "digital medical records") AND "nigeria" | 40 |
| AJOL | ("electronic medical records" OR "digital medical records") AND "nigeria" | 88 |
| Total |  | 294 |

**Table 2:** Summary of Included Studies.

| Authors | Study Purpose | Study Design | Study Location | Population | Outcomes measured | Findings |
| --- | --- | --- | --- | --- | --- | --- |
| Adereti 2019 [21] | To compare the quality of documentation using electronic- vs. paper-based SNCPs. | Longitudinal, quasi-experimental study | Public tertiary health facility in north central region with >500 bed capacity, 608 nursing care personnel | 32 nurses | Quality of documentation | Electronic-based SNCP:<br>Pre-test: 0%<br>1<sup>st</sup> post-test: 72%<br>2<sup>nd</sup> post-test, 54%<br>Paper-based SNCP:<br>Pre-test: 5%<br>1<sup>st</sup> post-test: 31%<br>2<sup>nd</sup> post-test: 32% |
| Aniekwe 2023 [24] | To assess the effect of implementing an electronic health information exchange (HIE) on the quality of HIV viral load data in Nigeria. | Longitudinal, quasi-experimental study | Nigeria, country-wide | 30 Healthcare facilities | Data completeness<br>Data validity or quality | Pre-test: 47%<br>6 months: 67%<br>*P*-value < 0.01<br>Pre-test: 90%<br>6 months: 91%<br>*P*-value < 0.01 |
| Hassan 2018 [23] | To assess patients' satisfaction with EMR at the hospital's pharmacy | Cross-sectional study | Federal Medical Centre, Keffi, Nassarawa State | 300 patients, >18 years; 142 male, 158 female | Satisfaction with pharmacy services received | 46% were satisfied with pharmacy services under EMR platform while 54% were not satisfied. |
| Kana 2019 [20] | To assess the feasibility of using an electronic growth monitoring system to monitor infant growth | Longitudinal Study | Child Welfare Clinic of Barau Dikko Teaching Hospital, Kaduna, Kaduna State | 2789 infant records | Completeness of Data collection | 89% had complete data; 68% had at least one follow-up visit; |
| Olagundoye 2021 [22] | To assess the effect of improving EMR using electronic diagnostic terminology instead of the manual coding process | Longitudinal, quasi-experimental study | 26 General hospitals in Lagos state | 52 clinical coders | Coding Accuracy or data quality | Pre-test: 76.3%<br>Post-test: 96.7% |

**Fig 1:**
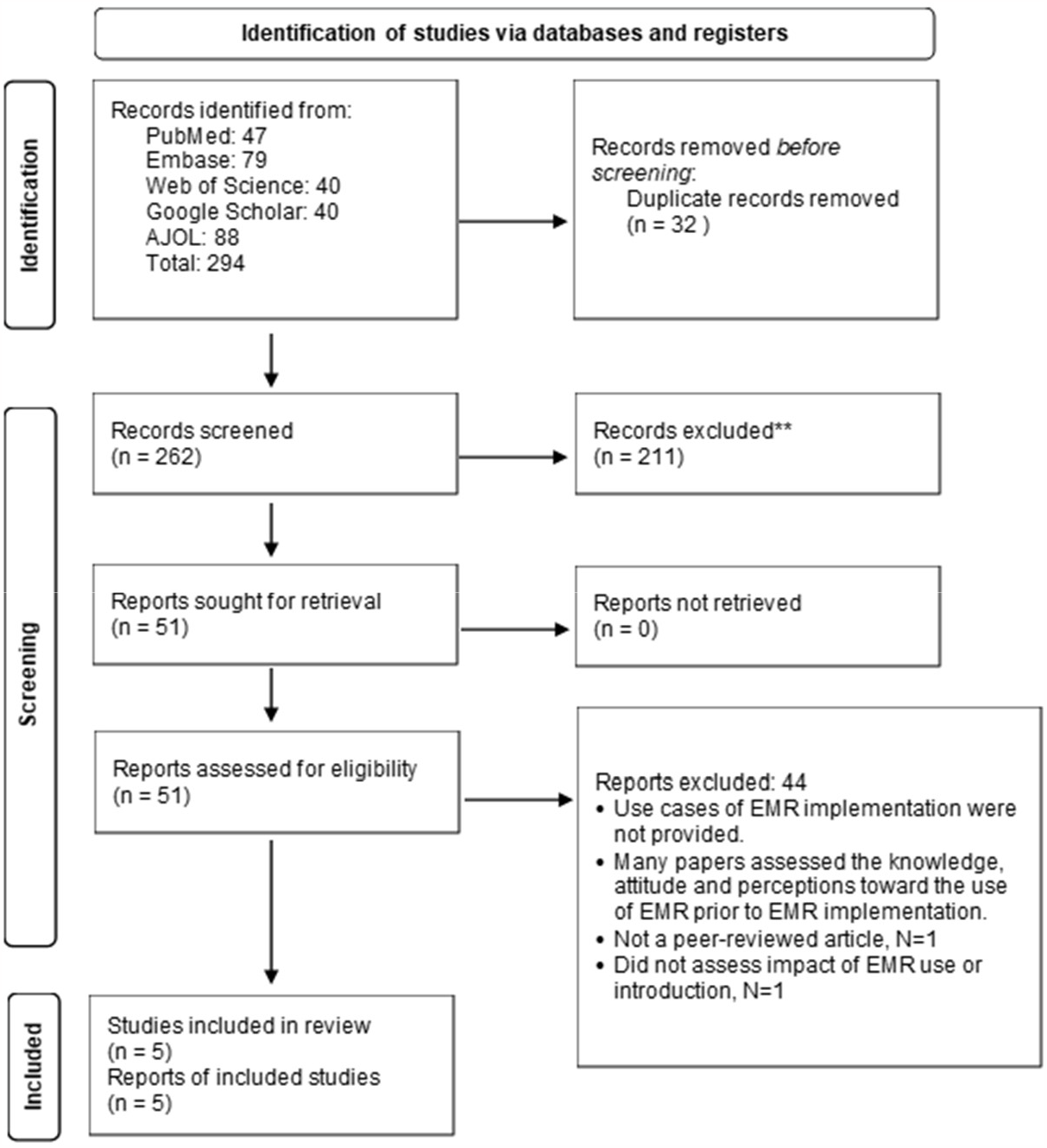
PRISMA flow diagram.

### Data quality

Four studies examined the influence of the introduction of [20,21,24] or improvements in [22] the EMR on data collection and documentation, with important implications for public health. The first, a longitudinal study [20], examined the feasibility of using the Child Electronic Growth Monitoring System (CEGROMS) to monitor the growth of infants in the Child Welfare Clinic of Barau Dikko Teaching Hospital, Kaduna, Kaduna State as part of the Kaduna Infant Development (KID) Birth Cohort Study [37] over nine months. The CEGROMS system included data capture forms, electronic tablets, and desktop computer linked to a cloud server. It collected data on child growth parameters such as head circumference, length, and birth weight. Participants were enrolled during birth or immunization visits and followed up during growth monitoring visits.

3,152 infants were assessed in this study. Of these, 89% had complete data and 68% had at least one follow-up visit. Data was more likely to be complete if infants were delivered at a health facility (OR=19.2; 95% CI: 13.7, 26.9), or were born to tertiary-educated mothers (OR=3.54; 95% CI: 2.69, 4.67). The odds of completing at least one follow-up visit were also greater for infants of tertiary-educated mothers (OR=1.33; 95% CI: 1.06, 1.51).

The second study [21] assessed the impact of utilizing electronic and paper-based Standardized Nursing Care Plans (SNCPs) on the quality of nurses’ documentation at a government-owned tertiary health facility. The design of the study was a pre-post quasi-experimental study comparing a new electronic SNCP to the existing paper-based SNCP. Whether hospital wards would use electronic or paper-based SNCP was determined by the hospital management. One adult medical wards using each of paper or electronic SNCPs was purposively selected for inclusion in the study. These wards had 32 nurses on staff and an average monthly admission of 34 patients.

Nursing documents for patients admitted for ≥ 4 days in these wards were assessed for pre-test (N=67), and first and second post-tests (N=68 each), 3 and 6 months later respectively. An instrument for assessing the quality of nursing diagnoses and documentation were used at each time point [38]. Following the introduction of SNCPs, there was a noticeable enhancement in documentation quality within both wards during the post-intervention period. While documentation quality in the electronic ward was good at 0% in the pretest, 72% in the first pretest and 54% in the second pretest, in the paper ward, it was 5% in the pretest, 31% in the first pretest and 32% in the second pretest, indicating a significant difference in the difference in documentation quality in both wards over time. This study underscores the crucial role of providing SNCPs in both electronic and paper formats in order to elevate the standard of nursing documentation.

The third study [24] assessed the impact of an automated Health Information Exchange (HIE) system on the validity and completeness of HIV viral load testing in healthcare facilities across Nigeria. The study design was longitudinal/quasi-experimental. The automated HIE system linked the EMR and the Laboratory Information Management Systems (LIMS) using APIs allowing the exchange of lab testing data. The validity of viral load data was assessed before the HIE was implemented and six months after. Three high-volume PCR labs and 30 treatment facilities were purposively selected for analysis.

The results indicated significant improvements in both data completeness and validity after the implementation of the automated HIE system. 15,226 records of specimens were analyzed at baseline and 18,022 records of specimens were analyzed six months later. Data completeness was defined as the percentage of observations with all the data regarding viral load testing, including the date and time the test was requested, specimen collected, tested, and so on. Data completeness increased significantly from 47% before HIE implementation to 67% six months after implementation (*P-*value <0.01). Data validity also increased from 90% before implementation to 91% after implementation (*P-*value <0.01).

The fourth study [22] was a longitudinal quality improvement (QI) study that aimed to use the Plan-Do-Study-Act cycle framework to enhance the accuracy of coding using the International Classification of Diseases, 10th revision (ICD-10) for morbidity/mortality data at general hospitals in Lagos State. The specific objective was to increase the accuracy rate from 78.7% to ≥ 95% between March 2018 and September 2018. Additionally, the study aimed to determine whether the electronic browsing tool could yield better coding accuracy compared to the traditional manual method. Thus, unlike the previous three studies that evaluated the introduction of EMR, this study studied improvements in the EMR.

The intervention involved introducing electronic diagnostic terminology software [39] and providing training to 52 clinical coders across the 26 general hospitals. The clinical coders were medical records personnel most of whom had a bachelor’s degree or its equivalent and had been in service a median of 7 years. To evaluate the impact of the intervention, an end-of-training coding exercise was conducted to compare accuracy between the old method and the new approach. Accuracy was defined based on the extent of correctly coded diagnoses. Over the period from April 2018 to March 2019, the accuracy rates in the hospitals increased from 76.3% to 96.7%, exceeding the QI goals.

The pooled percentage difference in data quality after introducing or improving the EMR was 123% (95% CI: 70% to 176%, *p-*value < 0.001) based on three studies of data validity [21,22,24]. The summary of the risk of bias and forest plot are shown in Figs 2 & 3 respectively. There was limited heterogeneity in the estimates (I^2^ = 0%, *p-*heterogeneity = 0.74). The funnel plot (Fig 4) was however symmetrical and the *p-*value for Egger’s test was 0.75. Meta-regression was not done.

**Fig 2.**
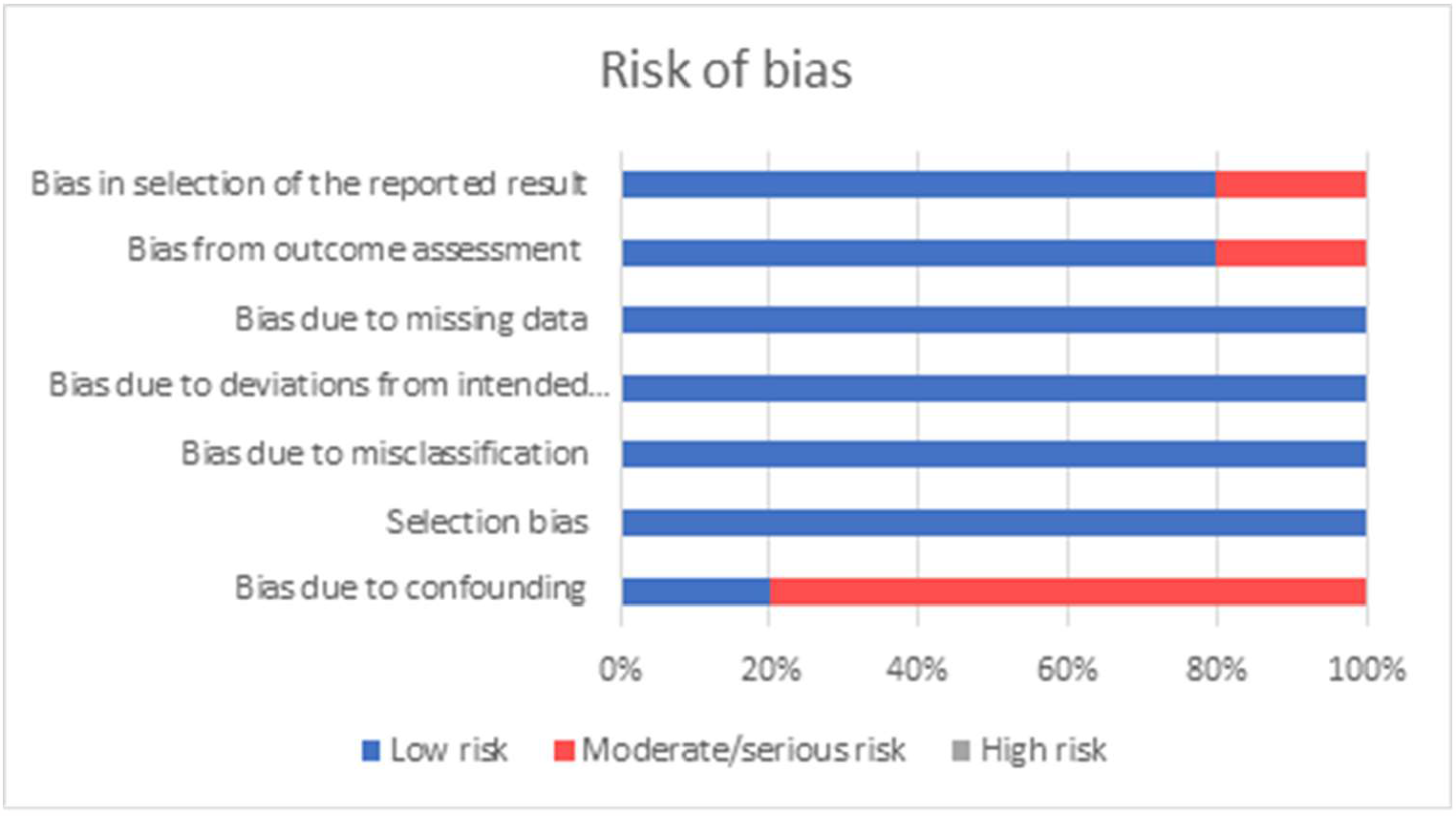
Summary of risk of bias.

**Fig 3.**
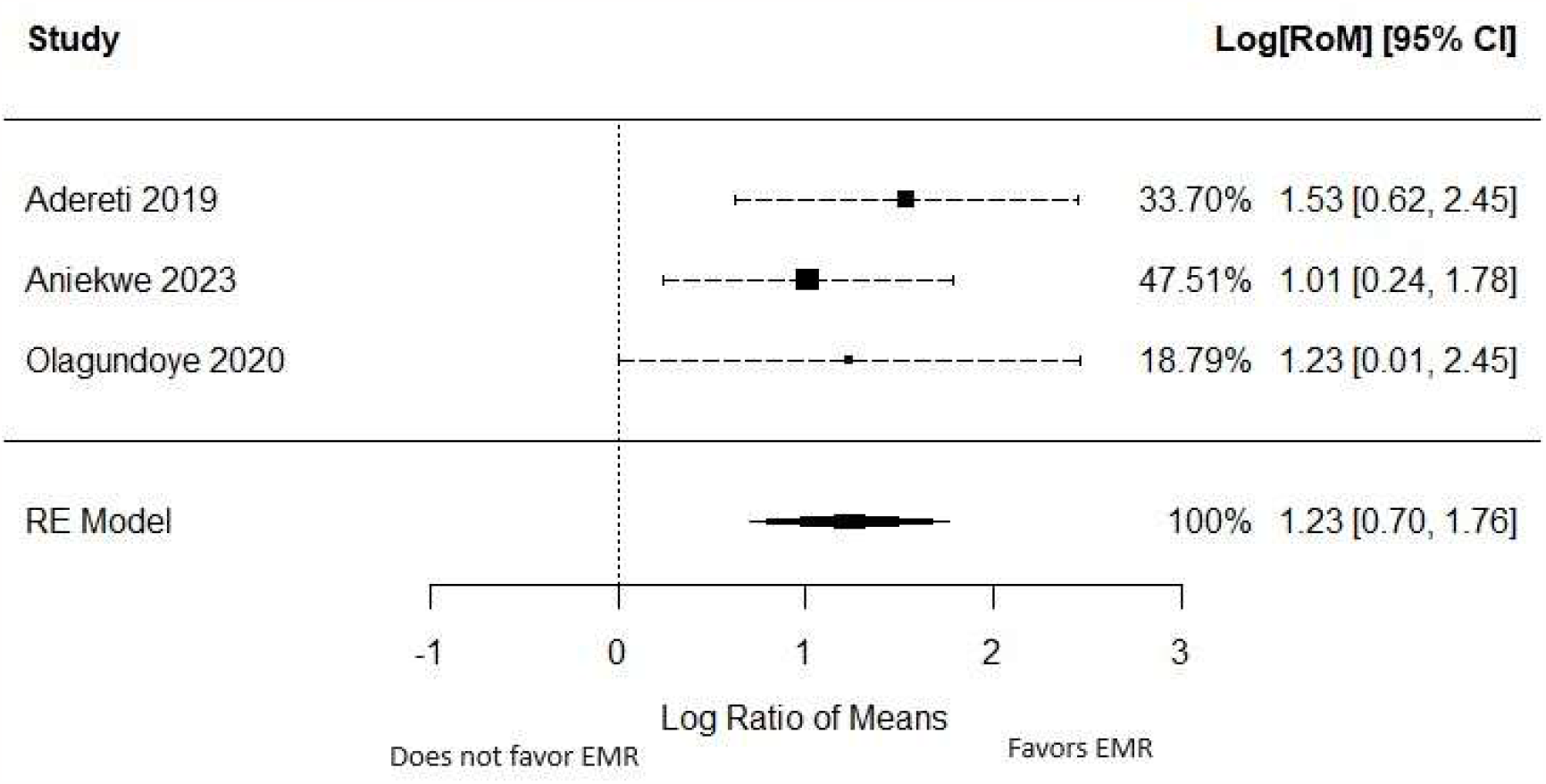
Forest plot of the effect of introducing EMR on data quality.

**Fig 4.**
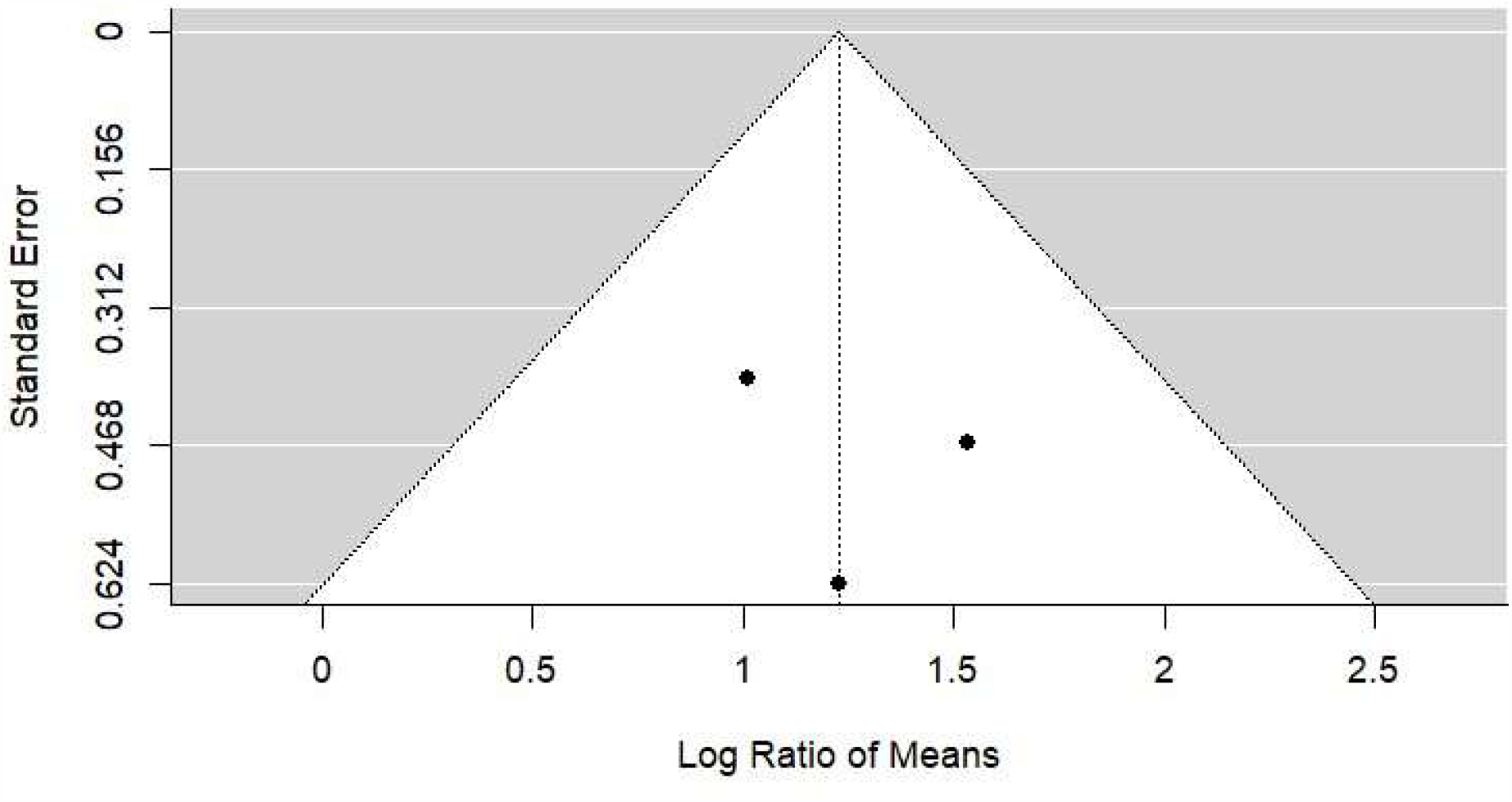
Funnel plot for the effect of introducing EMR on data quality.

Taken together, we conclude that the introduction of quality improvements to EMR enhanced data completeness and validity.

### Patient care and satisfaction

One study examined the impact of EMR on patient care and satisfaction [23]. The study had a cross-sectional design and was conducted among 300 patients at the Federal Medical Centre, Keffi, Niger State, with the aim of assessing patients’ satisfaction with EMR implementation at the hospital’s pharmacy. The participants (142 female, 158 male) were selected using a simple random sampling technique in which registered outpatients were the sampling frame. Using a questionnaire designed and validated by the investigators, the participants were asked about their satisfaction with the pharmacy services they accessed following the introduction of the EMR. 33% of the respondents believed that the EMR has improved services in the pharmacy department while 38% believed otherwise; the rest were indifferent. Only 28% were satisfied with the time it took to wait for service. Overall, 46% of the participants were satisfied. Unfortunately, waiting time was not objectively assessed in this study. The studied parameters were also not evaluated at baseline for comparison. The study did not also include a baseline assessment or a control group for comparison. We are therefore unable to conclude whether the introduction of the EMR improved patient satisfaction or shortened patient waiting times.

## Discussion

The evidence with respect to the impact of EMR use in Nigeria is very scant. We reviewed five primary studies that have examined the use and impact of EMRs in Nigeria. These studies assessed medical record quality and patient satisfaction following the introduction and use of EMR.

The EMR is central to the modernization of healthcare systems worldwide [40]. The transformative potential of EMRs lies in their ability to facilitate information management, streamline clinical workflows, and ultimately enhance patient care [41]. A striking facet that the studies unveil is the catalytic role of EMRs in mitigating errors resulting from incomplete or outdated information. The included studies have been examined in programs deploying EMR to strengthen public health programs [20], medical and nursing care [21,22] and laboratory testing [24]. The direction of our findings was similar – included studies consistently demonstrated that the EMR improved the quality of medical record data in Nigeria. We, thus, found that the introduction of the EMR improved data completeness by 123%.

Developed countries have seized the potential of EMRs to revolutionize healthcare by facilitating seamless data exchange, improving clinical decision-making, and enhancing patient engagement [42]. However, there remain disparities in EMR adoption, particularly in regions such as sub-Saharan Africa. Here, the challenges are manifold, encompassing not only technical hurdles but also socio-economic constraints and the digital divide in computer literacy among healthcare professionals [3]. These challenges are both emblematic of the complex digital landscape and unique to the local conditions of each developing country. For instance, it is incumbent upon any healthcare system to fortify its IT infrastructure to ensure seamless EMR operation. Ongoing training programs are also required to allow healthcare providers (HCPs) to operate the EMR applications with confidence and competence. There are also locally specific challenges such as epileptic power supply [43]. By fortifying IT infrastructure, fostering a culture of continuous learning, and providing robust support mechanisms, healthcare systems can navigate the intricacies of EMR implementation with a healthy amount of confidence.

Patient satisfaction, often deemed the bedrock of healthcare quality, reflects the synergy between medical expertise, operational efficiency, and the overall patient experience [44]. It is usually the result of reduced waiting times, enhanced information accessibility, and streamlined administrative processes. The transformation expected to be fostered by EMRs is not confined to operational efficiency; it extends to the very essence of healthcare—an essence deeply rooted in empathy, compassion, and patient-centricity [45]. HCPs would be afforded the time and tools to engage more closely with patients. The reduced administrative burden would pave the way for meaningful patient interactions, allowing HCPs to actively listen, educate, and engage with patients. It is noteworthy that HCPs in high-income countries believe that the EMR has actually increased administrative burden, though the actual impact on clinician burnout has not been rigorously studied [46]. Unfortunately, the reviewed studies did not rigorously evaluate the effect of introducing the EMR on patient satisfaction. Qualitative studies from Nigeria assessing patient and HCP satisfaction have had mixed outcomes [8,25], with the expectation that efficiency and satisfaction would improve as HCPs gained more familiarity with the EMR technology and the clinical workflow was improved. Studies from the USA, Kuwait and Saudi Arabia have reported improved patient satisfaction following the introduction of EMRs [47,48]. The rollout of EMRs in Nigeria is recent and improvements in patient satisfaction and healthcare quality can be expected as clinicians become more adept in the use of these tools.

This study has noteworthy strengths and limitations. By focusing on a single country, we are able to sharply focus on country-specific issues and potentially propose a national research agenda for digital health. Our meta-analysis of the effect of the introduction of the EMR on data quality could potentially be useful for health systems in the country aiming to assess the value of the EMR. Private facilities were however not included in any of the primary studies, though they account for a substantial proportion of healthcare in Nigeria [49,50]. The timeframe of the studies was also not long enough to evaluate any long-term impacts on data quality.

Going forward, additional research may improve our understanding of the value of the EMR in healthcare delivery. These include studies assessing health professionals’ readiness, impact on patient-relevant clinical outcomes in different clinical settings and a range of diagnoses, patient satisfaction using standard validated tools, objectively measured patient waiting time and quality indicators. It is important for the studies to carefully define the PICOS – participants, interventions, comparators, outcomes and study design [40]. It will also be important to ensure the applicability of collected data for clinical research and programs [51,52].

## Conclusion

We systematically reviewed the evidence linking the introduction of EMRs to data quality and patient satisfaction. We found that data quality increased substantially following the introduction of EMRs. We were unable to quantitatively assess the impact on patient satisfaction as there were too few studies and the included study did not have a rigorous design.

## Data Availability

N/A

## Acknowledgements

Nil

